# Suppression and Mitigation Strategies for Control of COVID-19 in New Zealand

**DOI:** 10.1101/2020.03.26.20044677

**Authors:** Alex James, Shaun C. Hendy, Michael J. Plank, Nicholas Steyn

## Abstract

A standard SEIR-type compartment model, parameterised for New Zealand, was used to simulate the spread of Covid19 in New Zealand and to test the effectiveness of various control strategies. Control aims can be broadly categorised as either suppression or mitigation. Suppression aims to keep cases to an absolute minimum for as long as possible. Mitigation aims to allow a controlled outbreak to occur, with the aim of preventing significant overloads on healthcare systems and gradually allowing the population to develop herd immunity.

Both types of strategy are fraught with uncertainty. Suppression strategies can succeed in delaying an outbreak, but only for as long as such control measures can be sustained. Once controls are eased or restricted, an epidemic is likely to follow as no herd immunity has been acquired. The success or failure of mitigation strategies can depend sensitively on the timing and efficacy of control measures, and require the ability to bring rapidly growing outbreaks under immediate control when needed. This is as yet untested even for a combination of national interventions including case isolation, household quarantine, population-wide social distancing and closure of schools and universities.

Although there are disadvantages to both types of approach, suppression has the advantage of buying time until a vaccine and/or treatment become available and allowing NZ to learn from rapidly unfolding events in other countries. A combination of successful suppression, strong border measures, and widespread contact tracing and testing resulting in containment could allow periods when control measures can be relaxed, but only if cases are reduced to a handful.

**Executive Summary:**

- Suppression strategies aim to keep the number of cases to an absolute minimum for as long as possible. This requires early and effective control interventions.
- Suppression can only delay an epidemic, not prevent it, but may buy enough time for a vaccine or treatment to become available.
- Mitigation strategies aim to control an epidemic so that herd immunity is acquired by the population without overwhelming healthcare systems.
- Mitigation strategies are likely to be very high risk: they are unproven internationally, potentially sensitive to uncertainty, and it may take years for herd immunity to be acquired.
- Strategy can be switched from suppression to mitigation. For example, once successful mitigation strategies have been tested in other countries. It is likely to be difficult or impossible to switch from a mitigation to a suppression strategy.
- A combination of successful suppression, strong border measures, and widespread contact tracing and testing resulting in containment could allow periods when control measures can be relaxed, but only if we can reduce cases to a handful.

## Introduction

The COVID-19 outbreak originated in Wuhan China in November 2019 before spreading globally to become a pandemic in March 2020. The human population currently lacks immunity to COVID-19, a viral zoonotic disease with a case fatality rate (CFR) of the order of 1%. This means that without controls there is likely to be widespread infection, which may overwhelm health care systems and lead to large numbers of deaths. In this study we examined potential control scenarios generated by a standard mathematical modelling approach to epidemic spread to consider the impacts on the progression of the disease in New Zealand. Our model is parameterised for the spread of COVID-19 through the New Zealand population (Wilson, 2020) with intervention strategies calibrated from a recent study of COVID-19 spread through the US and UK (Fergusson, 2020). Using the model we examine a range of possible interventions and their effect on the healthcare system and population fatality rate.

## Methods

We compare the outcomes of suppression and mitigation strategies in Covid-19 using a simple model with New Zealand specific parameters. The model is an ordinary differential equation model with susceptible (S), exposed (E), pre-symptomatic (P), infectious (I) and recovered (R) compartments. Cases are divided into untested (unconfirmed) infections (I_u_, R_u_) and confirmed cases (I_t_, R_t_) (Figure 1). The model is adapted from a model parameterised by Wilson et al (2020) for Covid19 spread in the NZ population (see Appendix for full model specification). Key assumptions include:

**Figure 1.**
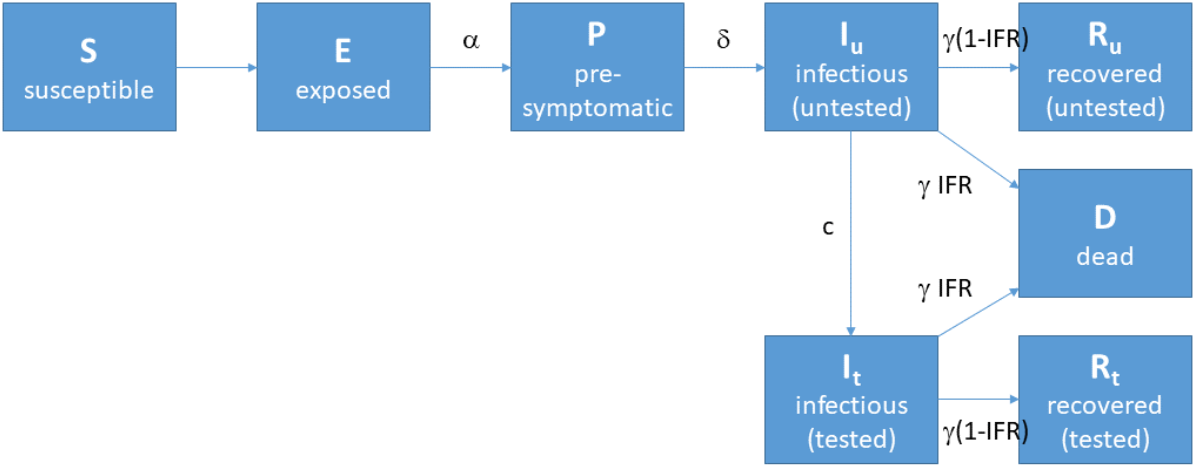
Diagram showing the compartment model used for Covid-19.

- A basic reproduction number of R_0_=2.5 (we also tested values of R_0_=3 and R_0_=2).
- Relative infectious in the pre-symptomatic period is 15% of that in the symptomatic period (Wilson et al. 2020), although it is possible this is an underestimate (Gayani et al, 2020).
- An overall infection fatality rate (IFR) of 1% provided the number of current ICU admissions is below hospital ICU capacity; when ICU is over capacity the excess infections have a 2% fatality rate. The IFR range of 1-2% spans the range estimated using age-specific fatality rates published by the Centre for Disease Control (CDC, 2020) combined with the age distribution of the New Zealand population (StatsNZ) – see Table 1. Wilson et al (2020) assumed an IFR of 0.9%.

**Table 1.**
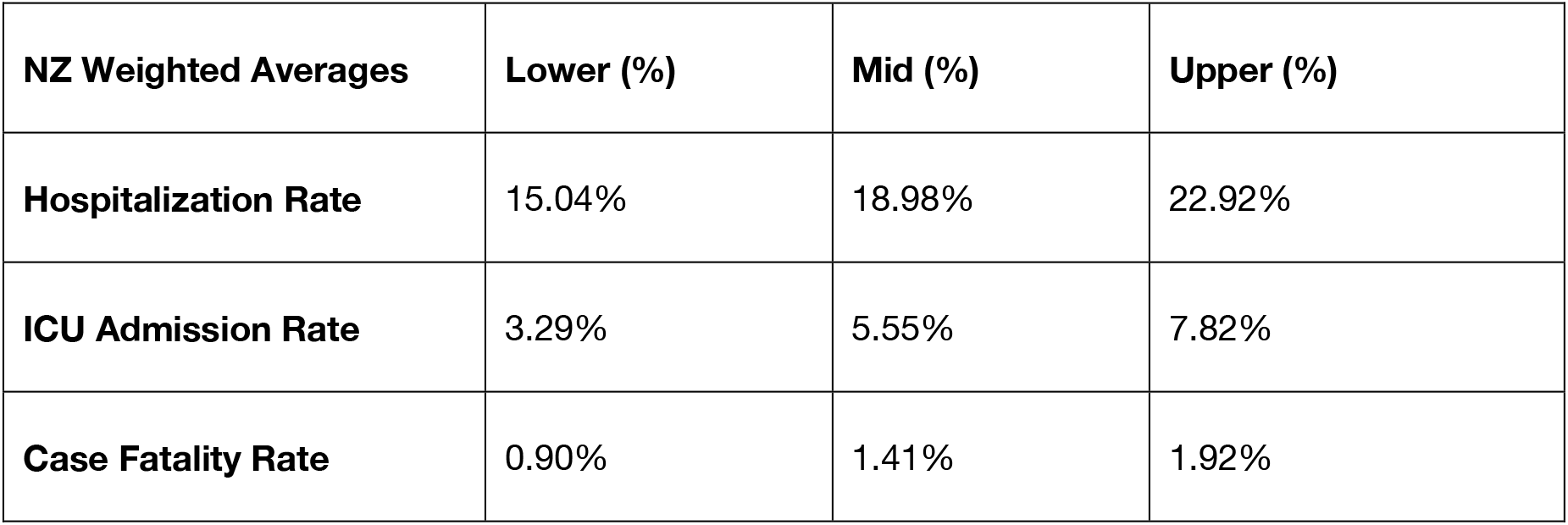
Estimates for the hospitalisation rate, ICU admission rate and case fatality rate (CFR) for New Zealand. These were calculated from the age-specific rates published by CDC (2020) combined with the age structure of New Zealand’s population (StatsNZ). These values assume that NZ has a similar healthcare system, rates of underlying health conditions, and Covid-19 testing rates as the US. Note that our model uses lower values than these estimates, which is reasonable because the model parameters represent rates per infection, which will be lower than rates per confirmed case if some infections are asymptomatic, subclinical or otherwise undiagnosed.
- A testing rate for symptomatic infections of 0.1 day^-1^. This is equivalent to assuming that 50% of infections are tested before either recovery or death and 50% of infections go untested. In reality, this ratio will vary depending on the number of current infections and the testing capacity and protocols, but most of our results are not sensitive to this assumption.
- Model simulations were initialised with 20 seed exposed infections introduced on 1 March 2020 (and no subsequent imported infections).

| NZ Weighted Averages | Lower (%) | Mid (%) | Upper (%) |
| --- | --- | --- | --- |
| Hospitalization Rate | 15.04% | 18.98% | 22.92% |
| ICU Admission Rate | 3.29% | 5.55% | 7.82% |
| Case Fatality Rate | 0.90% | 1.41% | 1.92% |

We measured hospital capacity in terms of the number of cases in ICU. We assumed that 5% of infections required hospitalisation, and of these 25% required ICU, meaning that overall 1.25% of all infections require ICU. These values are taken from Wilson et al (2020). These hospitalisation and ICU rates are substantially lower than estimates from CDC (2020) (see Table 1). However the latter could be biased by low detection rates in the US. In 2001, New Zealand had approximately 6 ICU beds per 100,000 people (Ministry of Health, 2005). We used a higher number of 10 ICU beds per 100,000 people here, to allow for conversion of other hospital facilities, for example from cancelling elective surgery, to temporary ICU beds. This gives a total of 500 ICU beds and so ICU capacity is reached when the total number of current infections reaches 40,000 (equivalent I = 0.8% of the population). We also investigated the consequences of having higher/lower ICU capacity.

We simulated two types of control strategy: suppression and mitigation. Both types of strategy were modelled by reduction in the transmission coefficient, resulting in a proportional reduction in the reproduction number with control (R_c_). This is a very simple control model which assumes that transmission rates from pre-symptomatic, unconfirmed symptomatic and confirmed symptomatic infections are all reduced by a constant factor. This describes society-wide control interventions, such as social distancing, hygiene measures, and lockdowns. It does not account for control measures that specifically target confirmed cases, such as case isolation. These could be modelled by a larger reduction in transmission rates for confirmed cases; we did not attempt this but it could be included in future model refinements to investigate the dependence of control efficacy on testing. The magnitude of the reduction in R_c_ was calibrated by comparisons with international data on case trajectories (Fig. 2) and modelling studies for the UK and US outbreaks (Ferguson et al 2020). The latter study assumed that interventions would have a similar impacts as they do with seasonal influenza. We then simulated suppression strategies by a fixed reduction in R_c_ for a period of 400 days. We simulated mitigation strategies by dynamic changes in R_c_ aimed at keeping the demand on the healthcare system under capacity (i.e. current infections under 40,000). In all cases, control began at t = 42 days after exposure of the initial seed infections.

**Figure 2.**
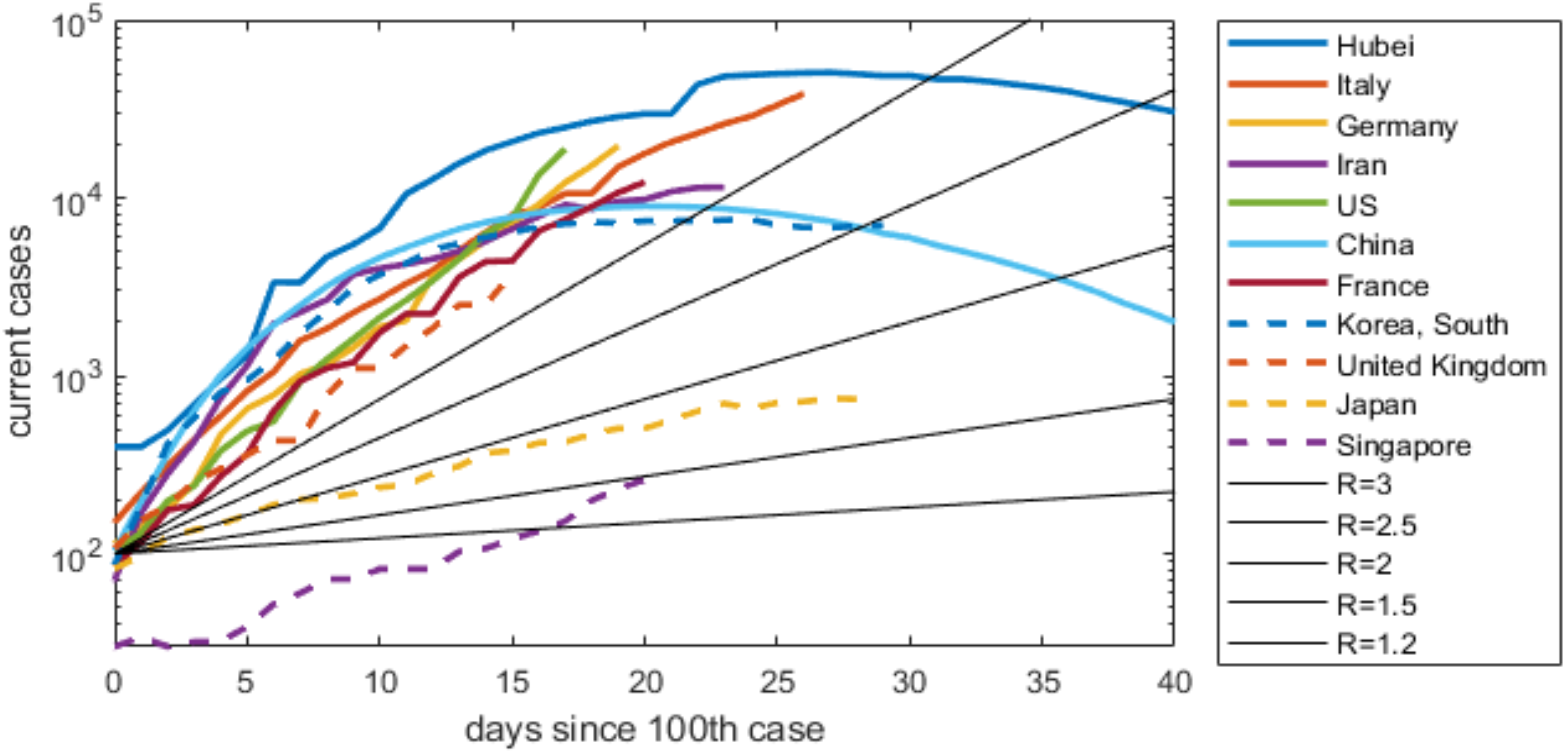
Number of current (non-recovered) cases as a function of time since the 100^th^ case, for 9 countries/regions with major outbreaks. China is split into Hubei province (blue) and the rest of China (purple). Data current as at 20 March 2020. Straight black lines show the expected gradient of the current cases trajectory when the effective reproduction number (number of new cases per existing case) is R = 1.2, 1.5, 2, 2.5, 3. Italy, France, Germany, UK and US are all close to an exponentially growing trajectory with R ≈ 3. Japan and Singapore have slower growth with R between 1.5 and 2. Countries on a downward trajectory (Hubei, rest of China, South Korea) have R<1.

## Results

### Suppression

We simulated an uncontrolled epidemic (R_c_=2.5) and five levels of increasingly strong control that reduce R_c_ to 2.3, 2, 1.75, 1.3, 1.2 (Fig. 3). These correspond approximately to the control measures modelled by Ferguson et al (2020) of: (i) closing schools and universities (R_c_=2.3); (ii) case isolation (R_c_=2); (iii) case isolation and household quarantine (R_c_=1.75); (iv) case isolation, household quarantine, and population-wide social distancing (R_c_=1.3); and (v) case isolation, household quarantine, population-wide social distancing, and closing schools and universities (R_c_=1.2).

**Figure 3:**
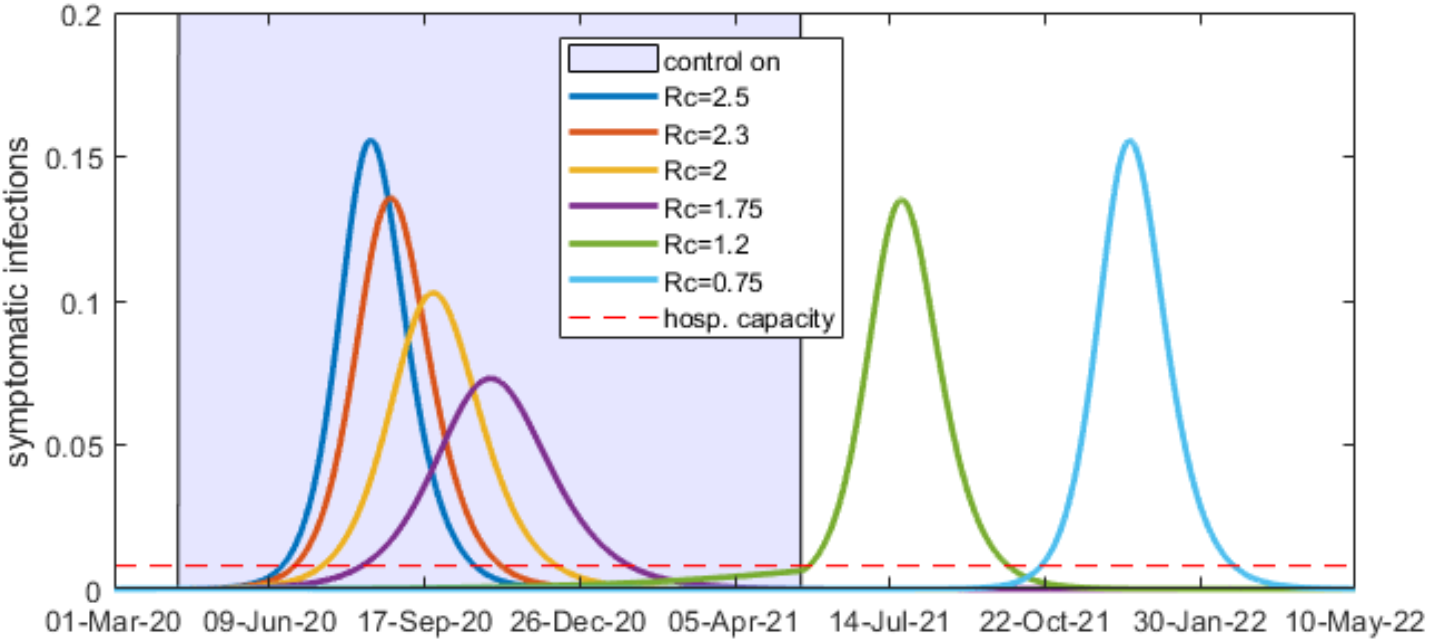
Supressing transmission for 400 days. At low levels of suppression the peak number of infections is still many times the effective hospital capacity but the outbreak is over within a year. At higher levels of suppression, the epidemic can be held in check for the 400-day control period, but a major outbreak occurs when control is lifted after 400 days. Any strategy with the final number of total infected less than about 60% will have a second wave of infections once controls are lifted. Dashed horizontal line shows hospital capacity.

However, note that there is considerable uncertainty and country-dependent variation in the effectiveness of any given control intervention, and these values of R_c_ should be seen as a range of potential outcomes, rather than a prediction of a given control measure.

An uncontrolled epidemic is expected to overwhelm hospital capacity by a factor of more than 10. Of the control scenarios, (i)-(iii) reduce the magnitude of the peak but do not succeed in suppressing the epidemic. Hospital capacity is still exceeded by a factor of at least 6. Scenarios (iv) and (v) successfully suppress the spread for the period of 400 days in which they were applied. However, when controls are lifted after 400 days, an outbreak occurs with a similar peak size as for an uncontrolled epidemic. In other words, these strategies can delay but not prevent the epidemic.

Table 2 shows the effect of each scenario on the size of the peak and the total number of infections and fatalities.

**Table 2:** Estimates of achieved transmission levels under different control scenarios. At the highest control levels, the epidemic is successful supressed and the peak number of infections is kept below hospital capacity, for the time period over which control can be sustained (assumed to be 400 days). Estimates were found by matching peak infections with those in the individual based model of Ferguson et al (2020).

| Strategy | $R_c$ (effective transmission rate under control) | Peak reduction relative to no control | Final number infected (when control ends) | Population mortality (when control ends) |
| --- | --- | --- | --- | --- |
| No control | 2.5 | - | 89% (89%) | 1.67% (1.67%) |
| Close schools and universities | 2.3 | 13% | 86% (86%) | 1.59% (1.59%) |
| Case isolation | 2.0 | 34% | 80% (80%) | 1.44% (1.44%) |
| Case isolation and household quarantine | 1.75 | 53% | 71% (71%) | 1.25% (1.25%) |
| Case isolation, household quarantine, and population-wide social distancing | 1.2 | 96% | 88% (6.8%) | 1.58% (0.06%) |
| All the above | 0.75 | >99% | 89% (0.04%) | 1.67% (0.0004%) |

### Mitigation

Mitigation strategies were simulated as a combination of a low level of control (case isolation and household quarantine, reducing R_c_ to 1.75) with periods of high control as required to try and keep the number of cases under hospital capacity. There are various ways this could be achieved. One approach is a “switching” strategy (Fig. 4) in which strong controls are imposed when hospital capacity is close to full (shaded blue areas in Fig. 4) and relaxed when there is spare hospital capacity available. The scenario shown in Fig. 4 requires the strong control periods to reduce R_c_ below 1 (R_c_ = 0.75 during periods in strong control in Fig. 4). Strong control is initiated when the number of ICU cases reaches capacity, and is lifted when the number of ICU cases drops to 50% of capacity. This is comparable to the scenario tested by Ferguson et al (2020), where weak control means R_c_=1.75 (representing case isolation and household quarantine in the Ferguson et al (2020) model) and strong control means R_c_=0.75 (representing the above measures plus population-wide social distancing in Ferguson et al (2020)). Periods of alternating strong/weak control are required to continue for approximately 750 days to prevent hospital capacity from being exceeded.

**Figure 4:**
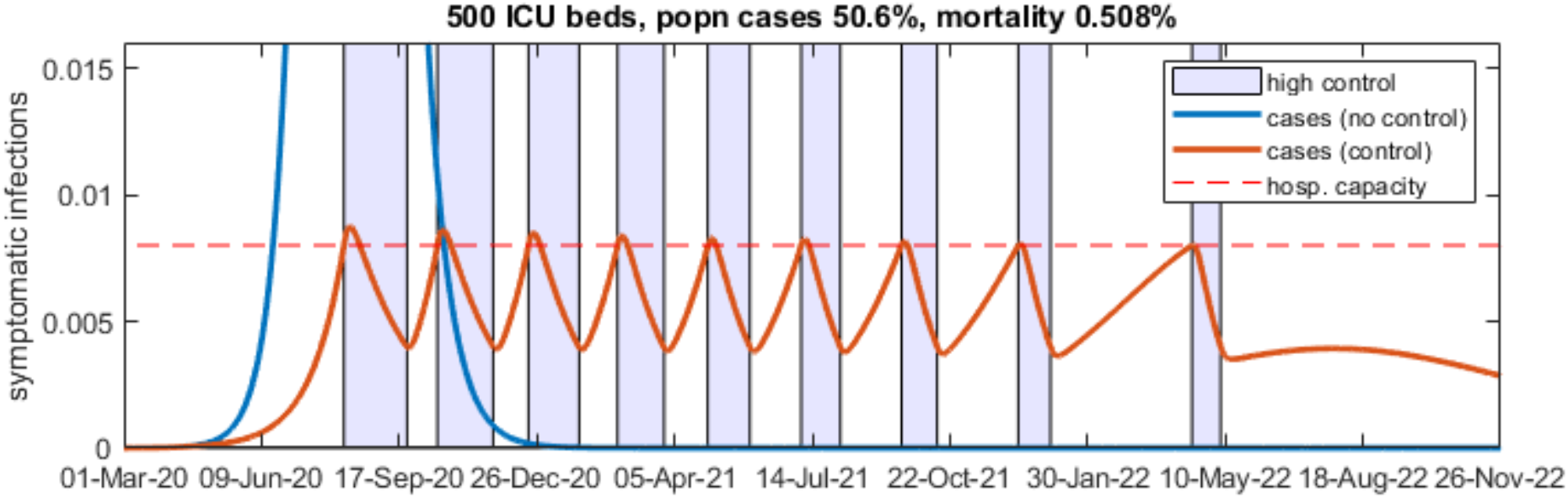
Mitigation strategies allow the disease to spread then apply control to reduce the peak allowing hospitals to be less overwhelmed. In general mitigation requires an initial period of weak control to allow the epidemic to establish, then an extended period (3-4 months) of very strong control. This can be followed by periods when control measures can be loosened, but strong control needs to be re-established when cases increase towards hospital capacity. One way to approach this is alternating periods of strong control (shaded blue, R_c_=0.75) and weak control (unshaded, R_c_ =1.75) as shown in the graphs.

**It is crucial to recognise that for the scenario shown in Fig. 4 to succeed requires control measures sufficiently strong to force R**_**c**_ **below 1. It remains completely unknown whether a scenario such as that shown in Fig. 4 is achievable in practice in New Zealand or any comparable country**. Fig. 2 shows that the only countries that have so far succeeded in getting the effective reproduction number less than 1 are China and South Korea. In those countries, this has been achieved by extremely intensive measures, including mandatory and strictly enforced quarantine, huge amounts of resources devoted to contact tracing, electronic surveillance of citizens’ movements, etc. In other countries, including those that have instigating major lockdowns such as Italy, there is as yet insufficient evidence that this has reduced Rc to 1 (Fig. 2).

We therefore tested the consequences of uncertainty in the effectiveness of strong control measures (Fig. 5), and in the timing of the trigger for initiating strong control (Fig. 6). If strong control only reduces R_c_ to 1.2, following the same strong/weak protocol as in Fig. 4 does not succeed in keeping (Fig. 5a), although it does still achieve a major reduction in peak size and overall mortality relative to the uncontrolled case. The outcome can be improved slightly by triggering strong control earlier (Fig. 5b, strong control initiated when ICU cases reach 50% of capacity). If strong control only reduces R_c_ to 1.5 (Fig. 5c), even with the earlier trigger, the mitigation strategy fails as hospital capacity is substantially exceeded and the population mortality increases to 1.0% (almost double that for Fig. 4).

**Figure 5.**
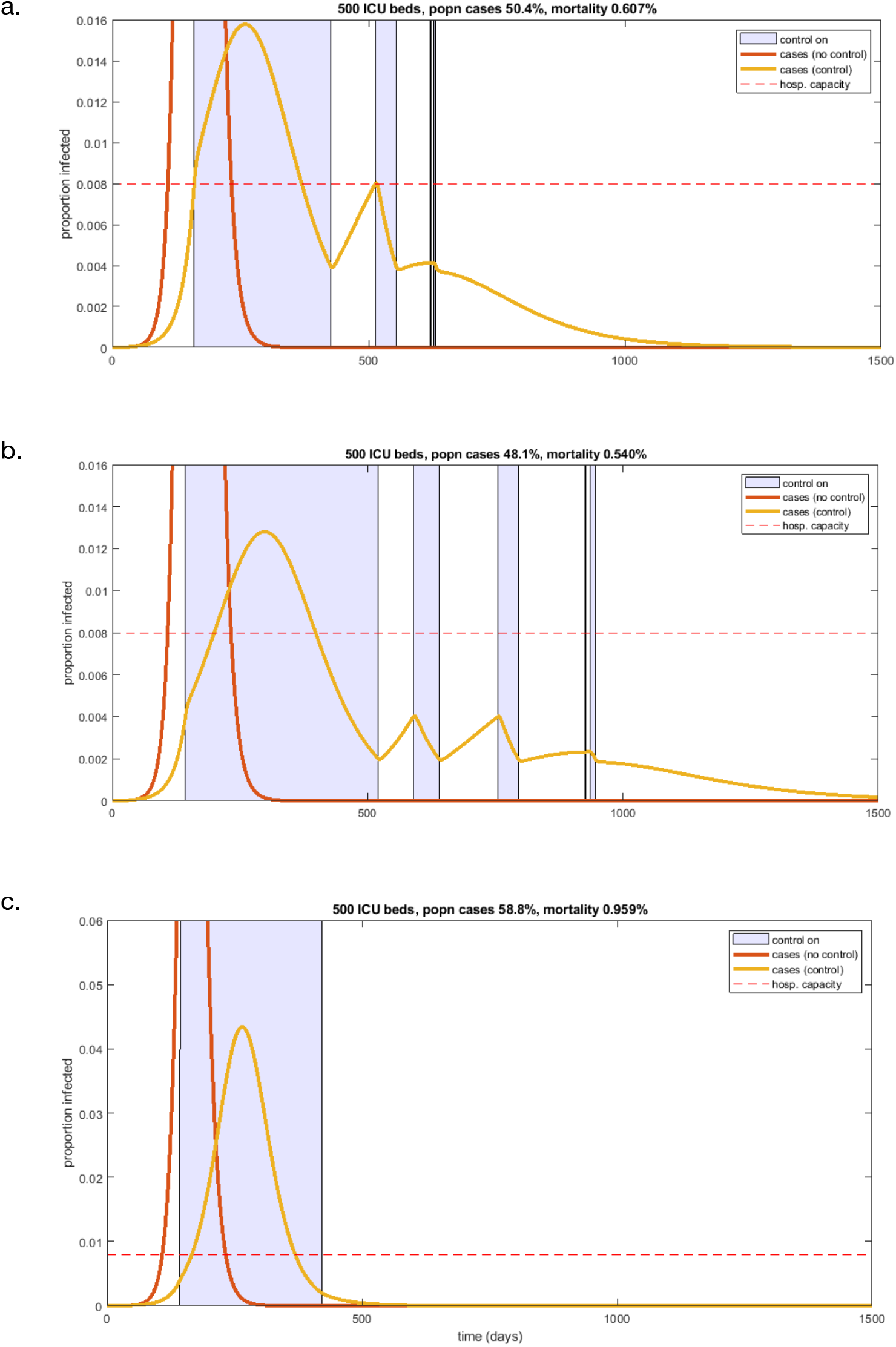
Mitigation strategies in which strong control (shaded blue time periods) fails to reduce the effective reproduction number R_c_ below 1. (a) Strong control is R_c_ = 1.2, trigger for strong control is when ICU reaches capacity; (b) Strong control is R_c_ = 1.2, trigger for strong control is when ICU reaches 50% capacity; (c) Strong control is R_c_ = 1.5, trigger for strong control is when ICU reaches 50% capacity. Weak control is (R_c_ =1.75) and the trigger for ending strong control is at 50% of the trigger for initiating strong control in all cases.

**Figure 6.**
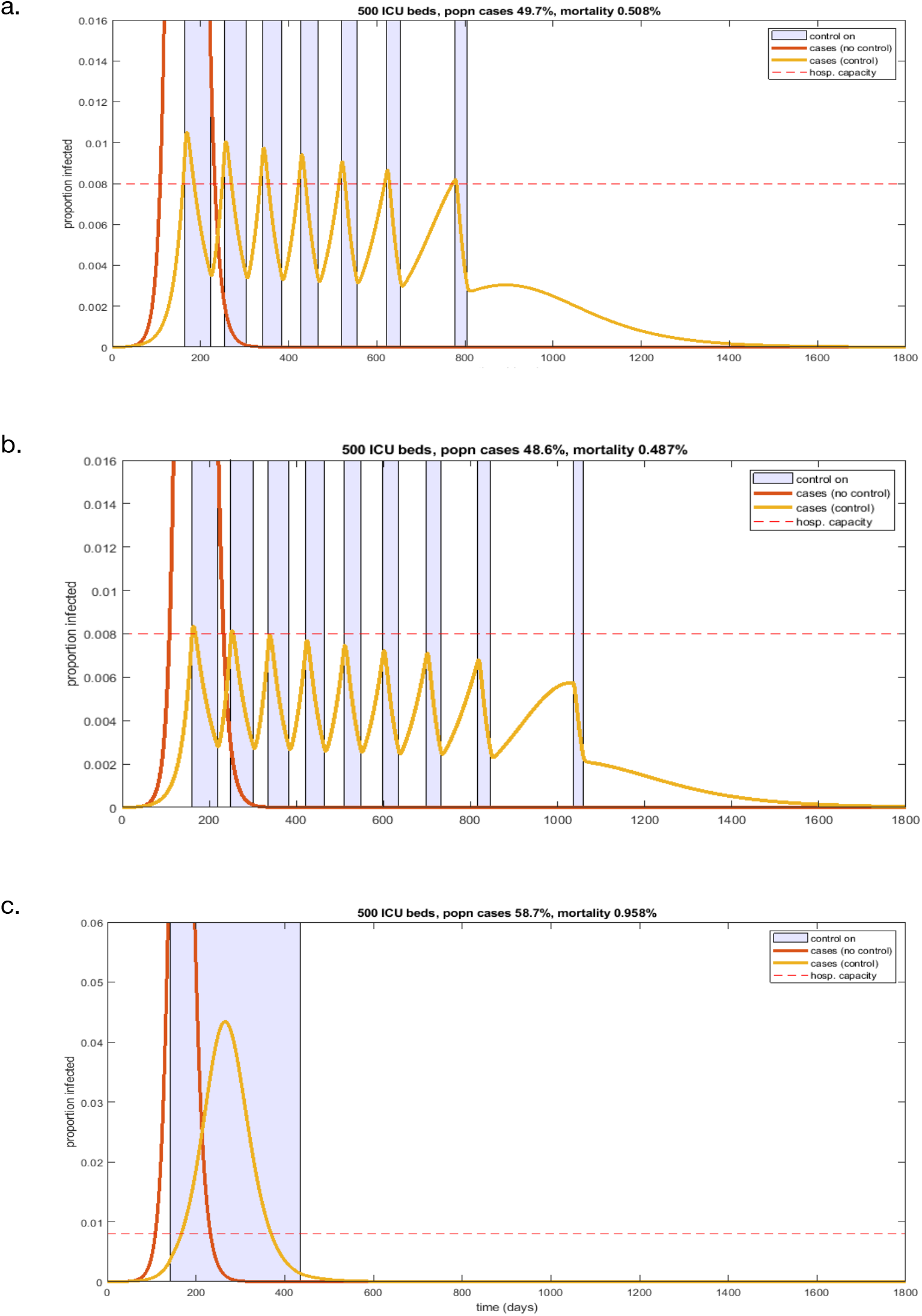
Mitigation strategies when the trigger for initiating strong control is the current number of confirmed cases, rather than the current total number of infections. (a) Strong control is R_c_ = 0.75, trigger for strong control is when ICU reaches capacity; (b) Strong control is R_c_ = 0.75, trigger for strong control is when ICU reaches 80% capacity; (c) Strong control is R_c_ = 1.5, trigger for strong control is when ICU reaches 40% capacity. Weak control is (R_c_ =1.75) and the trigger for ending strong control is at 50% of the trigger for initiating strong control in all cases.

The mitigation scenarios shown in Figs. 4-5 assume that a fixed proportion (1.25%) of all current infections are in ICU at any given time. In reality, there is a time lag between infections becoming symptomatic (compartment I_u_) and become severe enough to require ICU admission. To better reflect this, we assumed that no unconfirmed cases are in ICU and a fixed proportion of confirmed cases are in ICU. The proportion of confirmed cases in ICU was set at a level that gives the same overall proportion of ICU cases in the long run. The main consequence of this change to the model is that there is longer time lag before the strong control measures start to reduce the number of new infections, which means that cases overshoot hospital capacity (Fig. 6a). If strong control is sufficiently effective (R_c_=0.75), this problem can be offset by triggering strong earlier, when ICU reaches 50% of capacity rather than 100% of capacity (Fig. 6b). As before, if strong control is less effective than hoped for (only reducing R_c_ to 1.5),the mitigation strategy fails as hospital capacity is completely overwhelmed and the mortality rate doubles (Fig. 6c).

## Discussion

Mitigation strategies, which aim to allow the epidemic to go ahead at a controlled rate, keep demand on healthcare systems under capacity, and deliver herd immunity, are a tempting approach for the control of Covid-19. However, model results show that for these to be successful requires the ability to reduce transmission to a level where the effective reproduction number R_c_ is close to or below 1. It remains unknown whether this will be achievable in practice in New Zealand. There is no evidence that it has yet been achieved in comparable, western democracies, including those that have instigated major lockdowns such as Italy. The only regimes that have conclusively achieved this level of control are China and South Korea. In these countries, this has been achieved by extremely intensive measures, including mandatory and strictly enforced quarantine, huge amounts of resources devoted to contact tracing, electronic surveillance of citizens’ movements, etc. In addition, successful mitigation requires periods of these intensive control measures to be continued for up to 2.5 years before the population acquires a sufficient level of herd immunity. This could be an underestimate as these models do not include population turnover via birth-death, which will become significant over this time frame and may act to reduce the build-up of herd immunity. Furthermore, correct timing of strong control measures is crucial to successfully keeping healthcare systems from being overloaded. Small uncertainties in case trajectories could lead to drastically overshooting hospital and ICU capacity. If hospitalisation and/or ICU admission rates are in reality higher than assumed here, e.g. closer to the CDC (2020) estimates (Table 1), then mitigation strategies aimed at keeping ICU load under become even more difficult. Expanding New Zealand’s ICU capacity would alleviate this somewhat, and would be a sensible precaution in any case.

Suppression strategies aim to keep the number infections to a minimum for as long as possible, by early instigation of control measures. This alone cannot prevent an epidemic from taking place indefinitely because there is no acquisition of herd immunity. Once control is lifted, a serious outbreak is likely to take place. If control measures are lifted altogether, the eventual outbreak could be as serious as a completely uncontrolled epidemic, leading to population-wide mortality rates of around 2%. This mortality rate could be even higher if severe cases that cannot be treated because of hospital overload experience a significantly higher CFR.

A major advantage of suppression strategies as opposed to mitigation is that early suppression buys time. This has two key benefits: (1) it may be possible to delay the epidemic for long enough that a vaccine and/or effective treatment become widely available in NZ; and (2) it allows NZ to learn from rapidly unfolding events in other countries. This could include learning which mitigation strategies are most successful, and how to ensure timing of control interventions is robust to uncertainty.

The simulations in this study were initialised with 20 seed infections and assumed no subsequent arrivals of new infections from overseas. Significant numbers of imported infections could accelerate the spread in the early stages of epidemic. This could have important consequences for the timing of control interventions. A separate, forthcoming study will investigate the effects of restricting international and domestic flights one the epidemic trajectory.

If strong suppression is successful in reducing the number of cases close to zero, it is possible that some control measures could be lifted. This would require: (i) continued widespread testing and contact tracing to ensure there are no undetected case clusters; and (ii) strong border measures to remain in place to ensure no fresh infections are imported. This approach is similar in principle to the on-off strategy shown in Fig. 4, but with the crucial difference that it aims to keep cases close to zero (as opposed to merely under ICU capacity). As long as (i) and (ii) are in place and we are confident that there are no undetected cases, this could allow periods when schools, businesses and services can operate and many aspects of day-to-day life to continue.

## Data Availability

No new data is presented in this manuscript.

## Acknowledgements

The authors would like to thank Professor Mick Roberts from Massey University and Dr Lucy Telfar Barnard from the University of Otago for their useful comments on this work.

## Appendix

The appendix, containing the model specification, is available at www.tepunahamatatini.ac.nz.

